# The Epidemiology of Hundreds of Individuals Infected with Omicron BA.1 in Middle-Eastern Jordan

**DOI:** 10.1101/2022.01.23.22269442

**Authors:** Rima Hajjo, Mahmoud M. AbuAlSamen, Hamed M. Alzoubi, Raeda Alqutob

## Abstract

In less than two months of its detection in Jordan, lineage B.1.1.529 recognized as Omicron, is constituting 55% of all confirmed coronavirus disease of 2019 (COVID-19) infections causing a rise in the daily cases in the country. Herein, we report on 500 cases, among the first identified Omicron infections in Jordan. We also report on the genomic diversity of 25 Omicron viruses identified in nasopharyngeal swabs from Jordan. Our results indicated that 96% of study participants were vaccinated who had asymptomatic, mild or moderate disease. One unvaccinated individual developed severe disease. The median age of Omicron cases was 30 years, and most frequent disease symptoms were: fever, coughing, sore throat, runny nose, general fatigue and muscle/joint pain. Viral genomic analysis results revealed that the BA.1 is the dominant Omicron sublineage in Jordan, with 45 to 58 total mutations. We identified a few amino acid modifications that could impact the accuracy of some polymerase chain reaction (PCR) tests. In summary, infections caused by BA.1 seem milder than earlier infections. However, it is unknown whether this change is due to alterations in the immunity landscape of the infected population or is the result of viral genetic mutations that reduced viral virulence. Hence, comparing similar studies from different countries is likely to give us a get a better understanding of this variant, its behavior and the impact on disease characteristics.

## 1. Introduction

Almost two years after the declaration of the coronavirus disease of 2019 (COVID-19) pandemic by the World Health Organization (WHO), the disease-causing virus is still sweeping the globe, causing more fatalities, ravaging health care systems, and resulting in severe social and economic consequences. The high transmission rates of the virus across the globe are raising concerns for the constant emergency of new viral variants that can diminish herd immunity, cause new pandemic waves and/or allow the virus to become more deadly.

A new variant of concern (VOC) for the severe acute respiratory syndrome coronavirus 2 (SARS-CoV-2) has been recently identified in early November 2021. The variant has been designated as PANGO lineage B.1.1.529(1), and was given the Latin name Omicron by the World Health Organization (WHO)(2). B.1.1.529 has further diverged into three sub-lineages: BA.1 (the standard lineage), BA.2 and BA.3. Tracking BA.1 across different geographies is of paramount importance to get a better understanding of this new variant, and assess its impact on disease epidemiology and clinical outcome. Currently, Omicron-infections make up 55% of all confirmed COVID-19 cases in Jordan, causing a rise in the daily cases (Figure 1A).

**Figure 1.**
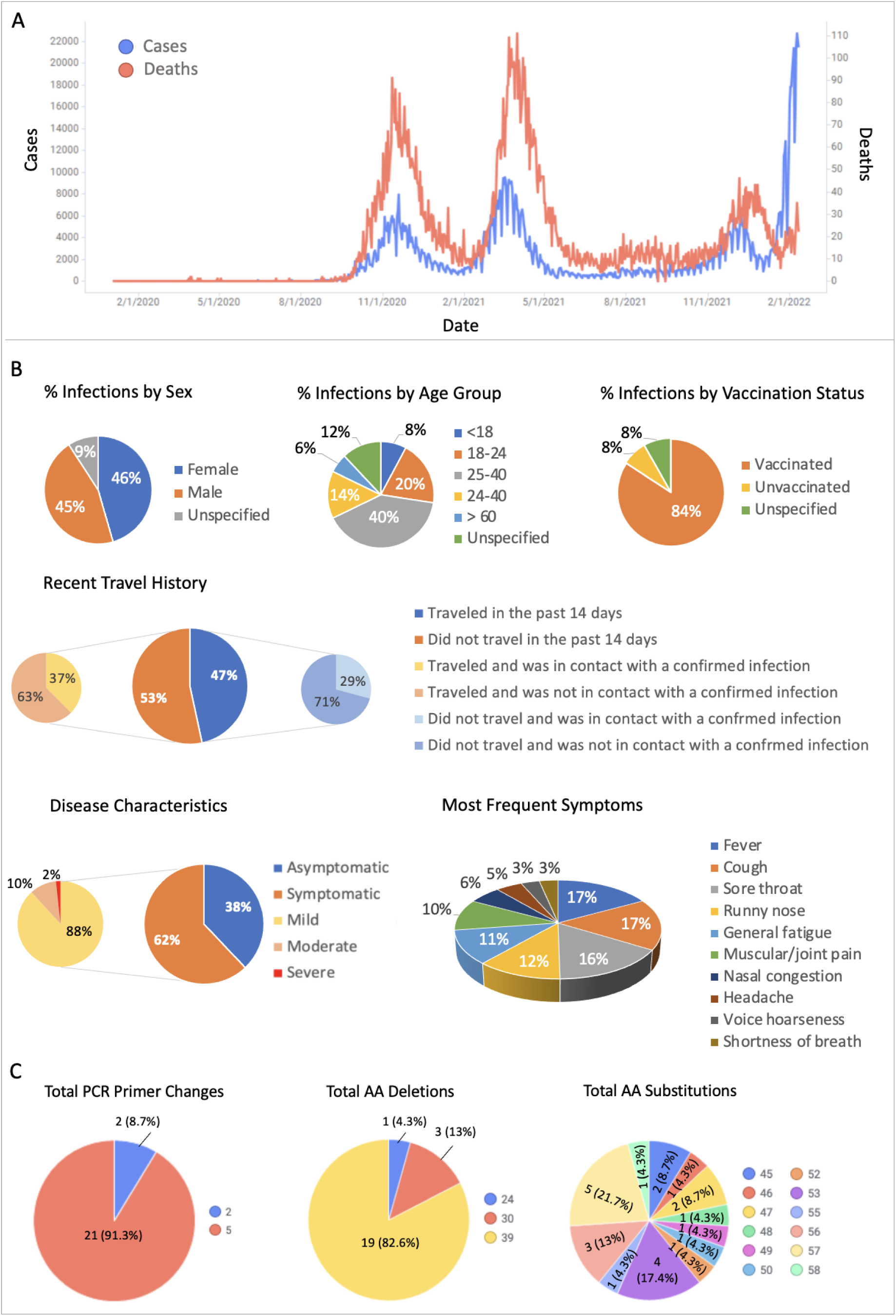
Epidemiologic and viral genomic data from Omicron cases in Jordan. A) Daily COVID-19 cases and deaths in Jordan over time as of February 12^th^, 2022. B) Summary statistics of epidemiologic data collected from 500 Omicron-infected cases from Jordan. C) Summary statistics of important mutational changes in Omicron viruses identified in Jordan and led to PCR primer changes and amino acid changes (deletions and substitutions).

In order to get a better understanding of Omicron BA.1, we sought to perform epidemiological description of the first identified Omicron infections to characterize these infections according to gender, age, vaccination status, prior SARS-CoV-2 infections, disease symptoms and symptom severity. We also characterized the viral genomic sequences downloaded from GISAID(3) for Omicron variants identified in Jordan. In fact, comparing similar studies from different countries around the world is likely to afford a better understanding of VOC BA.1, its behavior and the impact on disease characteristics across different geographies(4).

## 2. Materials and Methods

### 2.1. Epidemiologic Data

Jordan CDC has developed a questionnaire and shared it with collaborators at the Jordanian Ministry of Health to be used by contact tracing teams in order to collect the research data from Omicron-infected individuals described herein. Data collection by contact tracing teams was conducted using phone-based interviews with confirmed cases all over Jordan. As of January 4^th^, 2022, we collected contact tracing data for 500 cases. Textual data on symptoms were extracted, classified and combined into three levels of severity (mild, moderate and severe) according to Appendix Table 3. All statistics and analysis of proportions were performed in Microsoft^®^ Excel for Mac version 16.57. Some variables had missing data and was labeled through the manuscript and associated figure as “unspecified”.

A reinfection was defined as being reinfected after 90 days of a prior SARS-CoV-2 infection confirmed with a polymerase chain reaction (PCR) test. This study was approved by Al-Zaytoonah University Ethics Committee (IRB number: 29/11/2021-2022) and was conducted in accordance with the Declaration of Helsinki.

### 2.2. Sequencing Data

Sequencing specimens were collected using original nasopharyngeal swabs. Ion Torrent(5) assembly and Illumina MiSeq(6) were used as genome assembly methods depending on the originating lab. All sequencing specimens were collected by our collaborators at various diagnostic labs mentioned in the acknowledgement. Additional sequencing details are available on GISAID and GitHub: (https://github.com/rhajjo/JCDC_OmicronData).

### 2.3. Clade and Lineage Assignment

Nextclade in Bioconda version 1.9.0.0(7) has been used to identify mutations in comparison with SARS-CoV-2 reference sequence (WIV04/MN996528.1). Nexclade uses the identified mutations in order to assign the sequences to specific clades and to place them on a reference phylogenetic tree with a subset of all sequences available in GISAID(3).

### 2.4. Viral Genomic and Amino Acid (AA) Mutations

CoVsurver available from GISAID(3) was used to rapidly screen the Omicron genomes to screen AA changes in structural models and highlight if aa changes are close to common drug, host receptor or antibody binding sites.

## 3. Results and Discussion

### 3.1. Epidemiological Characterization of the First Complete Set of 500 SARS-CoV-2 Omicron Variant Cases in Jordan

Our results, which are based on reported data by 500 infected respondents (Table 1), revealed that 79.8% of the cases were reported in the capital Amman, while the rest of the cases were distributed among other largely-populated Jordanian governates including Balqa, Irbid and Zarqa. Results showed that 45.8% of all cases were males and 45.0% were females. The median age for the infected individuals was 30 years. Besides, 40.4% of the infections were in age group 25–40 years old (adults), 19.6% in age group 18-23 years old (youth), 7.8% in age group 2-17 years old (children) and 5.8% of all cases were in the age group 60 years old and above.

**Table 1.** Demographics data of study participants.

| Variable | Number | Percentage |
| --- | --- | --- |
| **Gender** |  |  |
| Female | 229 | 45.8 |
| Male | 225 | 45.0 |
| Unspecified | 46 | 9.2 |
| **Age** |  |  |
| <18 |  |  |
| 18-24 |  |  |
| 25 – 40 |  |  |
| 41 – 59 |  |  |
| > 60 |  |  |
| Unspecified |  |  |
| **Governate** |  |  |
| Amman | 399 | 79.8 |
| Irbid | 13 | 2.6 |
| Balqa | 12 | 2.4 |
| Zarqa | 7 | 1.4 |
| Madaba | 2 | 0.4 |
| Unspecified | 67 | 13.4 |

*Total sample size (N) = 500.*

**Table 2.** PCR primer changes in Omicron viral sequences identified in Jordan.

| **Nucleotide Change\*** | **Primer†** | **Number of Sequences‡** |
| --- | --- | --- |
| C26270T | Charité_E_F | 23 |
| C28311T | USCDC_N1_P | 23 |
| G28881A | ChinaCDC_N_F | 22 |
| G28882A | ChinaCDC_N_F | 22 |
| G28883C | ChinaCDC_N_F | 22 |

\*Observed nucleotide change in Omicron viral genomic sequences identified in Jordan by sequencing. †Impacted primers by the observed nucleotide change. ‡The number of viral genomic sequences from Jordan that contain the nucleotide changes listed in column 1.

Symptomatic infections varied in severity and constituted 51.4% of all Omicron infections among study respondents, while asymptomatic infections made up 31.4% of the total (the remaining 17.2% of the study sample didn’t have details on symptoms). Disease symptoms were mostly mild in 88.3% of the symptomatic infections, followed by moderate (9.7%) and severe symptoms (1.9%). The most frequent mild symptoms were fever (47.8%), coughing (47.1%), sore throat, (45.1%), runny nose (33.1%), joint and muscle pain (28.8%), general fatigue (31.5%), headache (13.2%), nasal congestion (16.3%) and hoarseness (9.3%). Notably, loss of taste and smell was only reported in 1.2% of the study cases.

Interestingly, 66.6% of the infected study individuals were fully vaccinated, *i.e*., had received their complete vaccine doses, and 14 days had already passed since their last dose. Disease symptoms were mainly mild in the fully-vaccinated group. However, individuals who were fully-vaccinated and received a booster shot within the last few months (*i.e*., 19.1% of the fully-vaccinated group) had asymptomatic infections in 44.1% of the cases, and mild symptoms in 45.5% of the cases, moderate symptoms in 3.9% of the cases, but none had severe disease.

Our results revealed that 8.6% of the study individuals were reinfected with the Omicron variant after 90 days from a prior SARS-CoV-2 infection. Most prior infections occurred between October 2020 and March 2021. Interestingly, reinfections ranged in severity from asymptomatic to severe, with the majority being either asymptomatic (41.9%) or mild (44.2%). Nevertheless, 2.3% of the reinfections were moderate, and 2.3% were severe. Epidemiologic results are summarized in Figure 1B. All details on the demographics data are available in Appendix (Appendix Tables 1-5).

### 3.2. Genomic Characterization of the First 25 Omicron Variant Viruses from Jordan

We analyzed all 25 complete viral genomes from Jordan, corresponding to Omicron viruses publicly available on the Global Initiative on Sharing All Influenza Data (GISAID)(8) database prior to January 15^th^, 2022. Viral genomics data were preprocessed according to the methods described by Rambaut *el al*(9), We retained 23 sequences, which had at least 95% coverage of the reference genome (WIV04/MN996528.1)(10) after trimming the 5′- and 3′-untranslated regions, and excluding sequences with > 5% ambiguous base calls (Ns). A maximum likelihood tree for 23 viruses was estimated, and viral lineages were defined by pangolin(1). DNA sequence variations (SNVs) on the nucleotide and amino acid levels were determined after performing pairwise alignments of the viral sequences with the reference genome WIV04(10) using CoVsurver enabled by GISAID (Appendix).

Omicron genomic sequences from Jordan were all assigned to sublineage BA.1 using Nextclade in Bioconda version 1.9.0.0(12). The analyzed sequences differed in the total number of mutations which ranged from 45 to 58, as shown in Appendix (Appendix Table 6). These mutations included amino acid (AA) deletions and AA substitutions that could impact the sensitivity of many PCR tests (Figure 1C). In fact, 21 (91.3%) sequences had 5 primer changes and 2 (8.7%) sequences had 3 primer changes in three types of PCR tests (Table 1). These primer changes have impacted the sensitivity of the corresponding PCR tests and obligated urgent updates on Omicron diagnostic tests. Detailed genomic analysis results are provided on GitHub (https://github.com/rhajjo/JCDC_OmicronData).

## 5. Conclusions

The observed changes in COVID-19 disease characteristics and Omicron’s genomic sequences highlight the need for large-scale studies that track the effects of viral mutational changes on disease epidemiology. Moreover, the identified positive impact of vaccines on reducing the severity of symptoms caused by Omicron infections could support the ongoing efforts to reduce vaccine hesitancy in Jordan and around the world(13).

## Data Availability

Raw data files and results of the genomic analyses are provided on GitHub (https://github.com/rhajjo/JCDC_OmicronData). The authors will make these data publicly available and supply additional details and a readme file upon request.

## Acknowledgments

We gratefully acknowledge the Jordanian Ministry of Health for providing the contact tracing data, in particular we extend our thanks to Dr. Ali Zaitawi, Head of Directorate on Communicable Diseases, and; Dr. Alaa Abu Hudaib, Director of Surveillance. We also acknowledge all the Authors from the Originating and submissting laboratories (Biolab Diagnostic Laboratories, Ministry of Health, Medlabs and Princess Haya Biotechnology Center) responsible for obtaining the specimens, performing the genetic sequencing experiments, reporting and saring the data via the GISAID Initiative (http://www.gisaid.org). RH acknowledges support from the Deanship of Scientific Research at Al-Zaytoonah University of Jordan (Grant number 2020-2019/17/03).

## Data Availability

Data files and results of the genomic analyses are provided on GitHub (https://github.com/rhajjo/JCDC_OmicronData). The authors will make these data publicly available and supply additional details and a readme file upon request.

## Supporting Information

Appendix for supplementary information on materials and methods, in addition to Appendix tables 1-6.

Appendix Table 1. Demographic data on Omicron-infected cases (n=500).

Appendix Table 2. Disease severity among Omicron-infected cases

Appendix Table 3. Frequency of disease symptoms in symptomatic Omicron infections. Appendix Table 4. Vaccination status among Omicron-infected cases.

Appendix Table 5. Reinfection statistics among Omicron-infected cases.

Appendix Table 6. List of mutations in 23 Omicron viruses from Jordan.

## Notes

The authors declare no competing financial interest.

