## Appendix for "The Epidemiology of Hundreds of Individuals Infected with Omicron BA.1 in Middle-Eastern Jordan"

### Supplementary Data and Methods

#### Epidemiologic Data and study design

Jordan CDC has developed a questionnaire and shared it with collaborators at the Jordanian Ministry of Health to be used by contact tracing teams in order to collect the research data from omicron-infected individuals described here. Data collection by contact tracing teams was conducted using phone-based interviews with confirmed cases. As of January 4th, 2022, 958 records were retrieved and manually curated (*i.e.*, checked for duplications and missing data). A total of 500 cases were finally approved data analysis purposes. Textual data on symptoms were extracted, classified and combined into three levels of severity (mild, moderate and severe) according to Appendix Table 3. A reinfection was defined as being reinfected after 90 days of a prior SARS-CoV-2 infection confirmed with a PCR test.

#### Sequencing

Sequencing specimens were collected using original nasopharyngeal swabs. Ion Torrent(1) assembly and Illumina MiSeq(2) were used as genome assembly methods depending on the originating lab. All sequencing details are available on GitHub: (https://github.com/rhajjo/JCDC_OmicronData).

### Clade and lineage assignation

Nextclade in Bioconda version 1.9.0.0(3) has been used to identify mutations in comparison with SARS-CoV-2 reference sequence (WIV04/MN996528.1). The Nexclade tool uses the identified mutations in order to assign the sequences to specific clades and to place them on a reference phylogenetic tree with a subset of all sequences available in GISAID(4).

### Viral genomic and amino acid (AA) mutations

CoVsurver available from GISAID(4) was used to rapidly screen the omicron genomes to screen AA changes in structural models and highlight if aa changes are close to common drug, host receptor or antibody binding sites.

### Data Availability

Virus sequences reported in this study are available from GISAID (All BA.1 viruses collected from Jordan prior to January 15th 2022). Additionally, the genomic data and analyzing scripts used in the study can be accessed in a GitHub repository:

(https://github.com/rhajjo/JCDC_OmicronData).

1. Mangul S, Caciula A, al Seesi S, Brinza D, Măndoiu I, Zelikovsky A. Transcriptome assembly and quantification from Ion Torrent RNA-Seq data. BMC Genomics [Internet]. 2014 Jul 14 [cited 2022 Jan 16];15(5):1–11. Available from: https://bmcgenomics.biomedcentral.com/articles/10.1186/1471-2164-15-S5-S7
2. MiSeq System | Focused power for targeted gene and small genome sequencing [Internet]. [cited 2022 Jan 16]. Available from: https://www.illumina.com/systems/sequencing-platforms/miseq.html

3. Package Recipe “nextclade” — Bioconda documentation [Internet]. [cited 2022 Jan 16]. Available from: https://bioconda.github.io/recipes/nextclade/README.html

4. Shu Y, McCauley J. GISAID: Global initiative on sharing all influenza data – from vision to reality. Eurosurveillance [Internet]. 2017 Mar 30 [cited 2021 Jan 28];22(13):30494. Available from: https://www.eurosurveillance.org/content/10.2807/1560-7917.ES.2017.22.13.30494

**Appendix Table 1.** Demographic data on omicron-infected cases (n=500).

| **Gender** | **Number (Percent)** |
|---|---|
| Male | 229 (45.8) |
| Females | 225 (45.0) |
| Unspecified | 46 (9.2) |
| **Governorate** | |
| Amman | 399 (79.8) |
| Irbid | 13 (2.6) |
| Balqa’a | 12 (2.4) |
| Zarqa’ | 7 (1.4) |
| Madaba | 2 (0.4) |
| Unspecified | 63 (12.6) |
| Jerash, Aqaba, Karak and Ajloun | 4 (0.8) |
| **Age** | |
| Mean ± SD (years) | 32.6 ± 14.3 |
| Median age (Interquartile range) | 30 (22.0 – 38.8) |
| <18 years | 39 (7.8) |
| 18-24 years | 98 (19.6) |
| 25 – 40 | 202 (40.4) |
| 41 – 59 | 72 (14.4) |
| >60 | 29 (5.8) |
| Unspecified | 60 (12.0) |

**Appendix Table 2.** Disease severity among Omicron-infected cases

| **Disease severity** | **Number (Percent)** |
|---|---|
| **Symptomatic** | |
| Yes | 257 (51.4) |
| No | 157 (31.4) |
| Unspecified | 86 (17.2) |
| **Severity of symptoms in symptomatic cases (n=257)** | |
| Mild | 227 (88.3) |
| Moderate | 25 (9.7) |
| Severe | 5 (1.9) |

**Appendix Table 3.** Frequency of disease symptoms in symptomatic omicron infections.

| **Severity of symptoms (n=257)** | Number (Percent) |
|---|---|
| **Mild symptoms** | |
| Fever | 123 (47.8) |
| Cough | 121 (47.1) |
| Muscular and joint pains | 74 (28.8) |
| Diarrhea | 4 (1.6) |
| Loss of taste and/ or smell | 3 (1.2) |
| Headache | 34 (13.2) |
| Chills | 4 (1.6) |
| Sore throat | 116 (45.1) |
| General fatigue | 81 (31.5) |
| Voice hoarseness | 24 (9.3) |
| Runny nose | 85 (33.1) |
| Nasal congestion | 42 (16.3) |
| Vomiting and /or nausea | 3 (1.2) |
| **Moderate symptoms** | |
| Shortness of breath | 24 (9.3) |
| Bronchitis | 1 (0.4) |
| **Severe symptoms** | |
| Pneumonia | 3 (1.2) |
| Hospitalization | 2 (0.8) |

**Appendix Table 4.** Vaccination status among omicron-infected cases.

| **Vaccination status** | **Number (Percent)** |
|---|---|
| Vaccinated | 421 (84.2) |
| Non-vaccinated | 38 (7.6) |
| Unspecified | 41 (8.2) |
| **Number of doses** | |
| Vaccinated (1 dose) | 19 (3.8) |
| Fully vaccinated (2 doses) | 403 (80.6) |
| Unspecified | 78 (15.6) |
| **Breakthrough infections within less than 14 days of last vaccine dose** | |
| Breakthrough infections | 333 (66.6) |
| Non-breakthrough infections | 14 (2.8) |
| Unspecified | 137 (27.4) |
| **Severity of symptoms among those who received a booster dose (n=77)** | |
| Asymptomatic | 34 (44.1) |
| Mild | 35 (45.5) |
| Moderate | 3 (3.9) |
| Severe | 0 (0.0) |
| Unspecified | 5 (6.5) |

**Appendix Table 5**. Reinfection statistics among omicron-infected cases.

| **Had SARS-CoV-2 Infection before 90 days of omicron infection** | **Number (Percent)** |
|---|---|
| Yes | 43 (8.6) |
| No | 413 (82.6) |
| Unspecified | 44 (8.8) |
| **Severity of symptoms among reinfected cases (n=43)** | |
| Asymptomatic | 18 (41.9) |
| Mild | 19 (44.2) |
| Moderate | 1 (2.3) |
| Severe | 1 (2.3) |
| Unspecified | 4 (9.3) |

**Appendix Table 6.** List of amino acid mutations in 23 omicron viruses from Jordan.

| Location | Mutation | Count* |
|---|---|---|
| Jordan | NSP4_T492I | 23 |
| Jordan | N_E31del | 23 |
| Jordan | N_G204R | 21 |
| Jordan | M_A63T | 23 |
| Jordan | Spike_V1264M | 1 |
| Jordan | N_P13L | 23 |
| Jordan | Spike_A67V | 20 |
| Jordan | Spike_Q954H | 22 |
| Jordan | Spike_K417N | 23 |
| Jordan | N_D343G | 10 |
| Jordan | NSP3_S1265del | 23 |
| Jordan | Spike_T478K | 10 |
| Jordan | NSP3_Y129H | 1 |
| Jordan | Spike_S477N | 9 |
| Jordan | NSP6_I189V | 23 |
| Jordan | Spike_N856K | 23 |
| Jordan | Spike_E484A | 12 |
| Jordan | Spike_G339D | 23 |
| Jordan | NSP16_L126F | 1 |
| Jordan | Spike_N440K | 23 |
| Jordan | Spike_Y144del | 18 |
| Jordan | Spike_N211del | 23 |
| Jordan | Spike_G496S | 2 |
| Jordan | Spike_Q493R | 12 |
| Jordan | Spike_T547K | 23 |
| Jordan | Spike_G446S | 23 |
| Jordan | Spike_S375F | 23 |
| Jordan | Spike_A701V | 3 |
| Jordan | Spike_R346K | 4 |

| | | |
|---|---|---|
| **Jordan** | Spike_D614G | 23 |
| **Jordan** | NSP3_K38R | 23 |
| **Jordan** | Spike_G142D | 18 |
| **Jordan** | Spike_S373P | 23 |
| **Jordan** | N_R32del | 23 |
| **Jordan** | N_R203K | 21 |
| **Jordan** | NSP6_S106del | 23 |
| **Jordan** | NSP6_L105del | 23 |
| **Jordan** | Spike_V70del | 22 |
| **Jordan** | NSP12_G44S | 1 |
| **Jordan** | NSP15_A92V | 9 |
| **Jordan** | NSP14_I42V | 23 |
| **Jordan** | Spike_F643L | 2 |
| **Jordan** | Spike_Y145del | 18 |
| **Jordan** | M_Q19E | 23 |
| **Jordan** | Spike_N764K | 23 |
| **Jordan** | Spike_ins214EPE | 23 |
| **Jordan** | NSP12_P323L | 23 |
| **Jordan** | Spike_N679K | 22 |
| **Jordan** | Spike_S371L | 23 |
| **Jordan** | Spike_V143del | 18 |
| **Jordan** | Spike_H69del | 22 |
| **Jordan** | NSP6_G107del | 23 |
| **Jordan** | Spike_Y505H | 18 |
| **Jordan** | N_S33del | 23 |
| **Jordan** | NS3_L106F | 10 |
| **Jordan** | NSP5_P132H | 23 |
| **Jordan** | M_D3G | 23 |
| **Jordan** | Spike_L981F | 21 |
| **Jordan** | E_T9I | 23 |
| **Jordan** | NSP3_L1266I | 23 |

| | | |
|---|---|---|
| **Jordan** | Spike_Q498R | 19 |
| **Jordan** | Spike_P681H | 22 |
| **Jordan** | NSP3_V1069I | 3 |
| **Jordan** | Spike_H655Y | 23 |
| **Jordan** | Spike_T95I | 18 |
| **Jordan** | NSP3_T725I | 1 |
| **Jordan** | NSP3_A1892T | 23 |
| **Jordan** | Spike_N501Y | 19 |
| **Jordan** | Spike_D796Y | 23 |
| **Jordan** | Spike_N969K | 21 |
| **Jordan** | Spike_L212I | 23 |

*The number of viral sequences (viruses) that has the mutation.
